# Effect of Digoxin on Interstage Outcomes following Stage I Palliation for Functionally Univentricular Hearts: A Systematic Review and Meta-Analysis

**DOI:** 10.1101/2022.05.04.22274522

**Authors:** Rohit S. Loomba, Jacqueline Rausa, Enrique Villarreal, Juan S. Farias, Saul Flores

## Abstract

**Objectives:** The goal of this systematic review and meta-analysis is to investigate the effects of digoxin on outcomes following stage I palliation for functionally univentricular hearts.

**Data Sources:** We conducted electronic searches of PubMed, Ovid and Cochrane.

**Study Selection:** Inclusion criteria included publication dates 1970–2018, children with functionally univentricular hearts between stage I and stage II palliation who received digoxin were compared to those who did not.

**Data Extraction:** We identified 148 unique citations; 5 full-text articles were included in the final review. Data from 4 studies was pooled for meta-analysis.

**Data Synthesis:** A total of 4 studies with 1,498 patients were included in the final analyses. Patient enrollment occurred between 2003 and 2013. A majority of patients were born full-term and approximately 25% were diagnosed with a syndrome. The most common cardiac diagnosis was hypoplastic left heart syndrome (70%). The most common initial surgical palliation was the Norwood procedure with a right ventricle to pulmonary artery conduit. The total amount of deaths was 121 (12 digoxin group versus 109 no digoxin group). The interstage mortality was reduced in the digoxin group [OR 0.25(95% CI 0.14 to 0.47)]. There was no statistically significant difference in the presence of arrhythmias or other complications.

**Conclusions:** This systematic review and meta-analysis suggests that digoxin significantly decreases interstage period mortality with a concurrent significant decrease in weight for age. The odds of arrhythmia or other complications are not significantly different with digoxin based on current data.

## INTRODUCTION

The palliation of functionally univentricular hearts frequently presents important challenges[1]. While there has been much knowledge gained through the building experience of such palliation, several questions still remain. The time between stage I and stage II palliation (interstage period) continues to be a time of particular high risk for children with functionally univentricular hearts[2]. During this period oxygen delivery is dependent on the delicate balance between the resistances of the pulmonary and systemic circuits and the resulting balance of blood flow to the pulmonary and systemic circuits[3, 4].

Advances in the monitoring and care of these patients during the interstage period have led to decreases in mortality during this time[5]. A number of medications may be used during the interstage period, with many not having much objective data to support their use routinely[6]. One medication that has been demonstrated by some studies to have benefit in the interstage period has been digoxin. There has been a number of studies examining the role of digoxin in children with functionally univentricular hearts during the interstage period[7-10]. Digoxin was initially demonstrated to decrease interstage period mortality by Oster and colleagues and Brown and colleagues[7, 11]. However, other studies have demonstrated conflicting findings with increased risk of complications and arrhythmias after digoxin administration. We, therefore, conducted a meta-analysis to investigate the effect of digoxin on outcomes in children with functionally univentricular hearts during the interstage period.

## METHODS

### Manuscript search and identification strategy

A systematic review of the literature was conducted to identify manuscripts describing the outcomes in children who received digoxin after undergoing congenital heart surgery. This was a newly conducted review with no previous review protocol being present.

Published manuscripts were identified by searching PubMed, OVID, and Cochrane databases from 1970 to 2018. The following search terms were used to query these databases: “digoxin”, “congenital heart disease”, “congenital heart surgery”, “palliation”. Only English language publications were reviewed and there was no restriction on the year of publication. Manuscripts were initially screened using the title and by reviewing the abstract. Full text of manuscript was retrieved for manuscripts felt to be pertinent to the review after the initial screen using the title and abstract. Manuscripts were deemed suitable for inclusion if they met the following requirements: patients with functionally univentricular hearts having received digoxin were compared to those who did not receive digoxin. Studies not meeting these criteria were excluded. Once full text manuscripts were obtained these were then reviewed by two of the authors (SF, RL). An assessment for quality, bias and any disparities in scoring was discussed. The Cochrane Handbook for Systematic Reviews was used for quality evaluation. Any discrepancies between authors was discussed and a resolution was achieved by group decision.

### Data extraction

Data regarding baseline patient characteristics and identified outcomes were extracted from the manuscripts identified for inclusion. Trial data was extracted using a data collection form. The extracted data was doubled checked to ensure the integrity of the extracted data. If no information was available for a particular outcome this was also recorded. Authors of included studies were not contacted for additional data.

### Bias analysis

Bias analysis was performed with specific attention paid to patient selection, intervention selection, endpoint inclusion, and result reporting of identified studies.

### Data analysis

Meta-analyses were characterized using Comprehensive Meta-Analysis Version 3.0 (Biostat, Englewood, NJ). Qualitative analyses consisted of characterizing the patient cohorts while quantitative analyses consist of pooled analyses of endpoints of interest. The endpoints of interest for quantitative analyses were mortality during the interstage period, need for heart transplant during the interstage period, and arrhythmia during the interstage period. Pooled analyses were conducted and the resulting forest plots generated by use of RevMan Version 5.3 (The Cochrane Collaboration, Copenhagen, Denmark, 2014).

## RESULTS

### Manuscript identification and characteristics

A total of 148 manuscripts were identified with 78 remaining after removal of duplicates. Abstracts for these 78 manuscripts were reviewed and a total of 5 had their full-text reviewed. A total of 4 studies were identified for final inclusion. Study review and inclusion is outlined in figure 1. These four studies consisted of data from 1,498 patients. Of these, 392 (26%) received digoxin while the remaining 1,106 did not. Studies enrolled patients between the years of 2003 and 2013. A majority of patients were born full-term and approximately 25% were diagnosed with a syndrome in the studies where this information was available (Table 1).

**Figure 1.** Flowchart Demonstrating the Search Strategy and Search Results for Published Manuscripts

**Figure 2.** Forest Plot Demonstrating Impact of Digoxin on Mortality

**Figure 3.** Forest Plot Demonstrating Impact of Digoxin on Arrhythmias

**Table 1.** Study Characteristics

Two of the studies described the cardiac lesions included[7, 9]. In both of these studies a majority of patients were diagnosed as having hypoplastic left heart syndrome (approximately 70%). Approximately 5% had double outlet right ventricle with left sided hypoplasia, 4% had double inlet left ventricle, and 4% had unbalanced atrioventricular septal defect (Table 2).

**Table 2.** Cardiac Lesions by Study

Stage 1 palliation consisted of a Norwood with right ventricle to pulmonary artery conduit in a majority of patients (approximately 55%). The next most common stage I palliation was a Norwood with Blalock-Taussig shunt which was done in approximately 35% of patients. A smaller proportion of patients underwent a hybrid or Damus-Kaye-Stansel with Blalock-Taussig shunt (Table 3).

**Table 3.** Cardiac Surgical Palliation by Study

### Mortality

Interstage mortality was found to be lower in the digoxin group (odds ratio 0.26, 95% confidence interval 0.14 to 0.47). A total of 121 (8%) patients died during the interstage period. Of those having received digoxin, 12 (3%) died during the interstage period compared to 109 (10%) of those not having received digoxin. Interrogation of the forest plot demonstrates that all studies showed a reduction in interstage mortality in the digoxin group, with 3 of these studies demonstrating a statistically significant difference.

### Arrhythmia

Two studies described the endpoint of arrhythmia in the interstage period with an overall tendency towards lower odds of arrhythmia in the digoxin group (odds ratio 0.84, 95% confidence interval 0.07 to 10.55) although this was not statistically significant[8, 10]. A total of 23 (4% of total patients with data for this endpoint) experience arrhythmia during the interstage period. Of those having received digoxin 9 (5%) experienced arrhythmia compared to 14 (4%) in those not having received digoxin. Interestingly, the forest plot demonstrates that the two source studies were split in the direction of effect, with Oster and colleagues demonstrating a tendency towards decreased arrhythmia in the digoxin group while Truong et al demonstrated a tendency towards increased arrhythmia in the digoxin group although neither individual finding was statistically significant.

### Heart transplant

Only Truong and colleagues described the endpoint of heart transplant and demonstrated a tendency towards lower odds of heart transplant in the interstage period (odds ratio 0.25, 95% confidence interval 0.01 to 4.90) although this was not statistically significant[10]. A total of 3 patients underwent heart transplantation (1% of total patients with data for this endpoint) with all 3 not having received digoxin.

### Change in weight for age

Truong and colleagues described change in weight for age in the interstage period with a more negative weight for age change in those in the digoxin group (mean difference -0.65, 95% confidence interval -0.70 to -0.60)[10]. Ghelani and colleagues also investigated weight for age in interstage patients although they did not present the data in respect to digoxin explicitly although they did comment that no medication imparted a significant effect on weight for age[9].

### Digoxin complications

Oster and colleagues described complications in the interstage period with a tendency towards increased odds of complications in the digoxin group (odds ratio 1.18, 95% confidence interval 0.75 to 1.88) although this was not statistically significant[8]. These were overall complications although the publication did break them down by type of complication with infectious complications being the most frequent. None of the other studies commented on complications explicitly and do not describe patients in whom digoxin needed to be discontinued for any reason.

## DISCUSSION

This systematic review demonstrates an overview of currently published data regarding the effects of digoxin in the interstage period. It appears that digoxin does mediate significant decrease in mortality. However, this may come at decreased weight gain[10]. The finding of the change in weight gain for age in this cohort requires further investigation. Truong and colleagues demonstrated a decrease in weight for age in both groups, although the decrease was significantly greater in those having received digoxin[10]. Ghelani and colleagues also explored the issue of weight gain for age and felt that digoxin did not have a direct effect in their patient cohort based on their analyses[9].

The effect digoxin imparts on the interstage period mortality has unknown mechanisms. It is possible that it is by augmenting cardiac function and, thus, cardiac output and oxygen delivery[12]. Data on cardiac function during the interstage period was not available in these studies at different timepoints for the two groups, therefore, it is difficult to comment on the role of function augmentation. While it is also possible that digoxin may be reducing the incidence of arrhythmias[13], the data on arrhythmia does not seem to indicate a reduction in arrhythmia is necessarily the driving factor for the decrease in the interstage period mortality.

It is important to note that the current data comes from patients with a variety of diagnoses within the spectrum of functionally univentricular hearts. Additionally, the data comes from patients who underwent one of a variety of procedures in the spectrum of stage I palliation. The limited number of studies here does not allow for any meaningful analyses of the impact of diagnoses or stage I palliation type on the influence of digoxin on changes in interstage period characteristics.

Based on the current published data the design of a definitive study can be proposed. Such a study would be multicenter in nature and would enroll children with functionally univentricular hearts with a variety of cardiac lesions having undergone a variety of procedures as the stage 1 palliation. Such a study would capture birth characteristics such as birth weight and gestational age, surgical characteristics such as cardiopulmonary bypass time, systemic functional ventricular data immediately prior to starting digoxin, degree of atrioventricular valve regurgitation immediately prior to starting digoxin, and weight immediately prior to starting digoxin. Endpoints of interest that should be captured are arrhythmias, digoxin toxicity, discontinuation of digoxin for any reason, heart transplant, and mortality. For those who survive to stage 2 palliation, weight at the time of stage 2 palliation, systemic functional ventricular data at the time of stage 2 palliation, and degree of atrioventricular valve regurgitation at the time of stage 2 palliation should be collected. This would allow for evaluation of efficacy using the endpoints of transplant, mortality and safety using the various other endpoints. Subset analyses could then be conducted based on cardiac lesion and procedure done for stage 2 palliation.

Some limitations were identified in the present study. All studies were retrospective, with many having limited capacity for adjustment, and thus a risk of selection bias and residual confounding. Also, studies included had a wide variation of cardiac diagnoses, as well as significant heterogeneity in outcomes, which limited our capacity for subgroup analysis in selected circumstances. Lastly, it is possible that some patients may have been represented in more than 1 study as some of the study times overlapped and may partially reduce the overall sample size. However, this systematic review and meta-analysis provide an overview of the current literature regarding digoxin administration in children with functionally univentricular hearts in the interstage period. It is strengthened by the use of a comprehensive search strategy, rigorous screening and eligibility criteria and by transparent reporting of our findings.

## Conclusion

Digoxin significantly decreases interstage mortality with a concurrent significant decrease in weight for age. The odds of arrhythmia or other complications are not significantly different with digoxin based on current data.

## Data Availability

All data produced in the present work are contained in the manuscript

## ACKNOWLEDGMENTS

None

## Author Contributions

Rohit S. Loomba, study design, data collection, data analysis, manuscript preparation, final approval of manuscript

Jacqueline Rausa, study design, critical review of manuscript, final approval of manuscript Saul Flores, study design, data collection, data analysis, manuscript preparation, final approval of manuscript

---

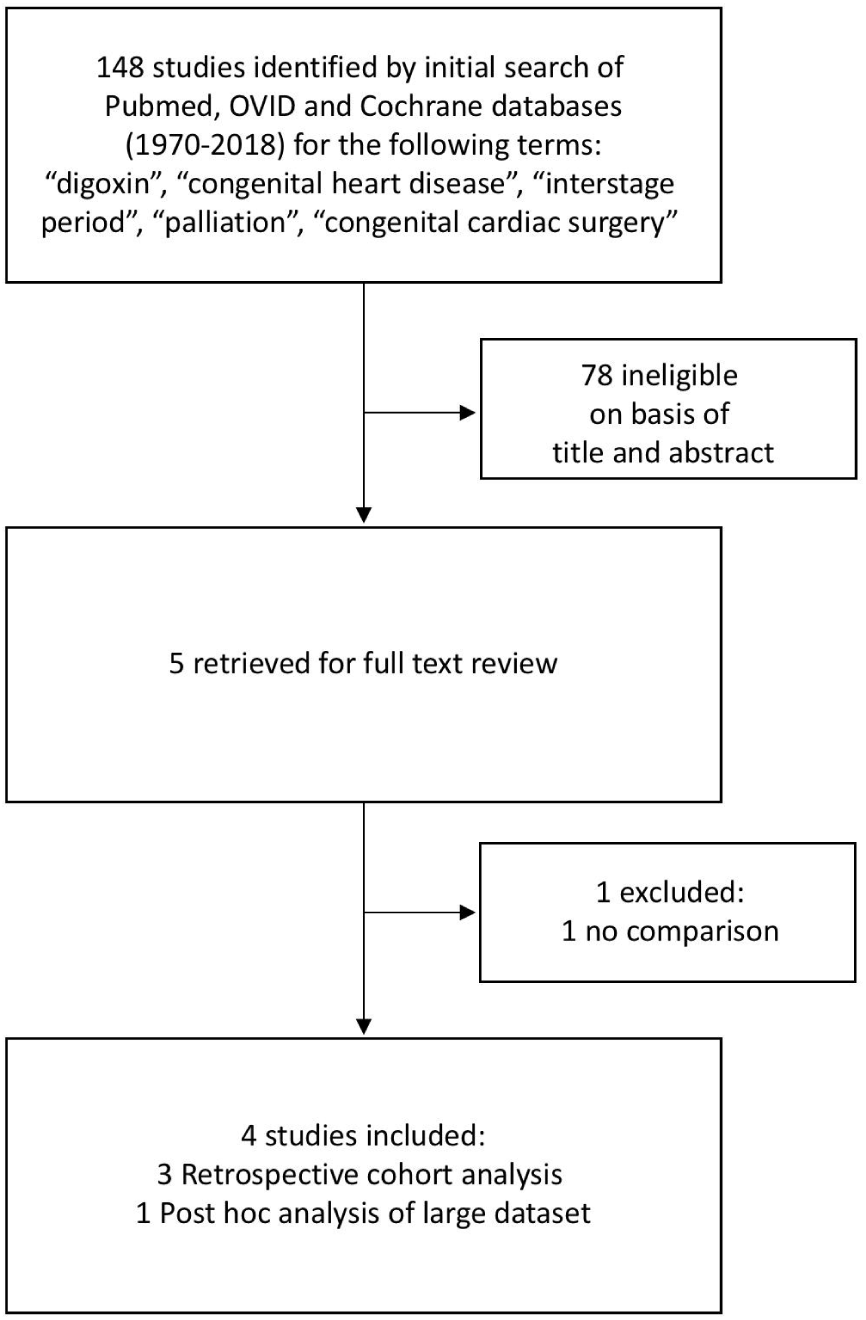

## REFERENCES

1. Ohye, R.G., et al., Comparison of shunt types in the Norwood procedure for single-ventricle lesions. N Engl J Med, 2010. 362(21): p. 1980–92.

2. Ghanayem, N.S., et al., Interstage mortality after the Norwood procedure: Results of the multicenter Single Ventricle Reconstruction trial. Journal of Thoracic & Cardiovascular Surgery, 2012. 144(4): p. 896–906.

3. Alsoufi, B., et al., Results of palliation with an initial modified Blalock-Taussig shunt in neonates with single ventricle anomalies associated with restrictive pulmonary blood flow. Ann Thorac Surg, 2015. 99(5): p. 1639-46; discussion 1646-7.

4. Ricci, M., et al., Near-infrared spectroscopy to monitor cerebral oxygen saturation in single-ventricle physiology. J Thorac Cardiovasc Surg, 2006. 131(2): p. 395–402.

5. Ohye, R.G., et al., Cause, timing, and location of death in the Single Ventricle Reconstruction trial. Journal of Thoracic & Cardiovascular Surgery, 2012. 144(4): p. 907-14.

6. Moffett, B.S., et al., Impact of pharmacotherapy on interstage outcomes in single ventricle infants. Congenit Heart Dis, 2011. 6(4): p. 286–93.

7. Brown, D.W., et al., Digoxin Use Is Associated With Reduced Interstage Mortality in Patients With No History of Arrhythmia After Stage I Palliation for Single Ventricle Heart Disease. J Am Heart Assoc, 2016. 5(1).

8. Oster, M.E., et al., Association of Digoxin With Interstage Mortality: Results From the Pediatric Heart Network Single Ventricle Reconstruction Trial Public Use Dataset. Journal of the American Heart Association, 2016. 5(1): p. 13.

9. Ghelani, S.J., et al., Impact of pharmacotherapy on interstage mortality and weight gain in children with single ventricle. Congenit Heart Dis, 2013. 8(3): p. 219–27.

10. Truong, D.T., et al., Digoxin Use in Infants with Single Ventricle Physiology: Secondary Analysis of the Pediatric Heart Network Infant Single Ventricle Trial Public Use Dataset. Pediatr Cardiol, 2018.

11. Oster, M., et al., Use of digoxin is associated with reduced interstage mortality: Results from the pediatric heart network single ventricle reconstruction trial. Circulation, 2015. 132.

12. McMahon, W.S., et al., Cellular basis for improved left ventricular pump function after digoxin therapy in experimental left ventricular failure. J Am Coll Cardiol, 1996. 28(2): p. 495–505.

13. Moffett, B.S., et al., Efficacy of digoxin in comparison with propranolol for treatment of infant supraventricular tachycardia: analysis of a large, national database. Cardiol Young, 2015. 25(6): p. 1080–5.

