## Supplementary material for "Effect of Digoxin on Interstage Outcomes following Stage I Palliation for Functionally Univentricular Hearts: A Systematic Review and Meta-Analysis": Table 1

| **Study** | **Birth weight (kg)** | **Gestational age (weeks)** | **Syndrome or genetic anomaly** | **Moderate or greater atrioventricular valve regurgitation** | **Moderate or greater depression of systemic ventricular function** | **Feeding tube during interstage period** |
| --- | --- | --- | --- | --- | --- | --- |
| ***Brown et al***  ***Digoxin (n=119)***  ***No digoxin (n=425)*** | 3.2 (entire cohort) | 39 (entire cohort) | 9 (8%)  32 (8%) | 20 (17%)  70 (16%) | 5 (4%)  11 (3%) | 66 (55%)  256 (60%) |
| ***Ghelani et al***  ***Digoxin (n=89)***  ***No digoxin (n=306)*** | --  -- | --  -- | --  -- | --  -- | --  -- | --  -- |
| ***Oster et al***  ***Digoxin (n=102)***  ***No digoxin (n=228)*** | 3.07  3.20 | 39  38 | 26 (31%)  52 (31%) | 22 (22%)  44 (19%) | --  -- | 21 (21%)  53 (23%) |
| ***Truong et al***  ***Digoxin (n=82)***  ***No digoxin (n=147)*** | --  -- | 38  38 | --  -- | --  -- | 17 (21%)  30 (20%) | 49 (60%)  70 (49%) |
