## Supplementary material for "Effect of Digoxin on Interstage Outcomes following Stage I Palliation for Functionally Univentricular Hearts: A Systematic Review and Meta-Analysis": Table 2

| **Study** | **Hypoplastic left heart syndrome** | **Double outlet right ventricle with left sided hypoplasia** | **Double inlet left ventricle** | **Double inlet right ventricle** | **Unbalanced atrioventricular septal defect** | **Other/unknown** |
| --- | --- | --- | --- | --- | --- | --- |
| ***Brown et al (n=544)*** | 369 (68%) | 23 (4%) | 26 (5%) | 1 (<1%) | 23 (4%) | 85 (15%) |
| ***Ghelani et al***  ***(n=395)*** | 290 (73%) | 25 (6%) | 14 (4%) | 0 (0%) | 15 (4%) | 51 (12%) |
| ***Oster et al***  ***(n=330)*** | -- | -- | -- | -- | -- | -- |
| ***Truong et al (n=229)*** | -- | -- | -- | -- | -- | -- |
