## Supplementary material for "Effect of Digoxin on Interstage Outcomes following Stage I Palliation for Functionally Univentricular Hearts: A Systematic Review and Meta-Analysis": Table 3

| **Study** | **Norwood with BT shunt** | **Norwood with RV to PA conduit** | **Hybrid** | **DKS with BT shunt** | **Other/unknown** |
| --- | --- | --- | --- | --- | --- |
| ***Brown et al (n=544)*** | 188 (35%) | 285 (53%) | 51 (9%) | 18 (3%) | 0 (0%) |
| ***Ghelani et al***  ***(n=395)*** | 121 (30%) | 227 (58%) | 31 (8%) | 12 (3%) | 4 (1%) |
| ***Oster et al (n=330)*** | 146 (44%) | 184 (56%) | -- | -- | -- |
| ***Truong et al (n=229)*** | 106 (46%) | | -- | -- | 123 (54%) |
